# Quantifying Cancer Clinical Trial Eligibility Using Artificial Intelligence-Based Matching

**DOI:** 10.64898/2026.06.03.26354859

**Authors:** Keshav Goel, Nathaniel J Myall, James Dickerson, Jennifer L. Caswell-Jin, Tyler Johnson, John Emmett Worth, Michael Gensheimer

## Abstract

**PURPOSE:** To develop and validate an artificial intelligence-enabled platform that combines efficient screening with explicit eligibility logic by converting unstructured cancer trial criteria into reusable structured queries, and illustrates potential eligibility using synthetic patients.

**METHODS:** We downloaded actively recruiting US interventional treatment trials for advanced/metastatic breast cancer, colon cancer, and non-small cell lung cancer from ClinicalTrials.gov. Medical oncologists created 24 synthetic patient vignettes. A large language model converted trial eligibility criteria into Structured Query Language (SQL) code and patient information into structured records, enabling rule-based matching without further LLM inference. Cancer details and treatment history were considered, but not laboratory results or comorbidities. Validation included physician editing of generated eligibility code for 30 trials and independent assessments by two physicians, followed by consensus adjudication, for eligibility across ten trials. We descriptively summarized potential eligibility by trial characteristics and when varying age, ECOG performance status, sex, and ZIP code.

**RESULTS:** Of 833 candidate trials, 746 met inclusion criteria. Performing matching for all 746 trials took a median of 0.459 seconds per patient, excluding patient-data preparation. Against physician consensus, automated matching agreed on 230/240 patient-trial pairs (95.8%; sensitivity 83.3%, specificity 100%). Across synthetic patients, potentially eligible trials ranged from 10 to 258 without geographic restrictions. Counts decreased with worse performance status and geographic restriction and were relatively stable across ages 30–90. Later-phase, randomized, and molecularly selective trials generally matched fewer vignettes.

**CONCLUSION:** LLM-generated SQL enables rapid, explicit rule-based screening for potential trial eligibility. Evaluation in synthetic vignettes supports feasibility, while missed matches underscore the need for further validation.

## Introduction

Despite widespread interest in cancer clinical trials among patients and oncologists, only around 7% of cancer patients enroll in therapeutic trials.^1^ Restrictive eligibility criteria, for instance based on performance status, contribute to this gap, and professional societies have been working to broaden these criteria.^2–4^ Geographic disparities also limit access, with clinical trial sites concentrated in urban and academic centers.^5–7^ Furthermore, a fundamental question remains unanswered: for a given cancer patient, how many trials are they actually eligible for, and how does this vary by patient characteristics? Answering this question at scale has been infeasible, as it would require manually matching detailed patient information against thousands of trial eligibility criteria, a process that is labor-intensive and error-prone.

Large language models (LLMs) have shown promise in automating eligibility determination for clinical trials.^8–12^ Several approaches use LLMs to assess individual eligibility criteria for each patient-trial pair, sometimes after an initial retrieval step. Repeating this inference across many patients and trials can be computationally demanding. We developed a methodology that uses LLMs to convert unstructured eligibility criteria into reusable Structured Query Language (SQL) queries. By converting patient information into structured form, we can efficiently check many patients’ eligibility for hundreds of trials. This separates the computational cost of interpreting trial criteria from repeated execution of explicit eligibility logic across patient records. We implemented this system in TrialFetch, a publicly available, non-commercial web application for doctors, research coordinators, and patients to find matching trials for a given patient.^13^

To demonstrate the utility of this methodology, we created synthetic patients with breast cancer, colon cancer, or non-small cell lung cancer (NSCLC) and matched them against actively recruiting advanced/metastatic trials for these cancers using the TrialFetch platform. We validated performance by comparing to manual determinations. We then summarized potential eligibility by trial characteristics and systematically varied vignette characteristics to illustrate changes in matching results. This work establishes a platform for future research on trial access and disparities.

## Methods

### Trial Database Construction

We developed a text-to-SQL pipeline that converts unstructured eligibility criteria and patient information into structured data for precise matching. This approach uses LLMs to parse free-text eligibility criteria into SQL SELECT statements, which can then be executed against a patient database to determine eligibility at scale. The pipeline converts both the eligibility criteria of each trial and the patient’s medical information into structured data that can be precisely matched against each other (Figure 1).

**Figure 1.**
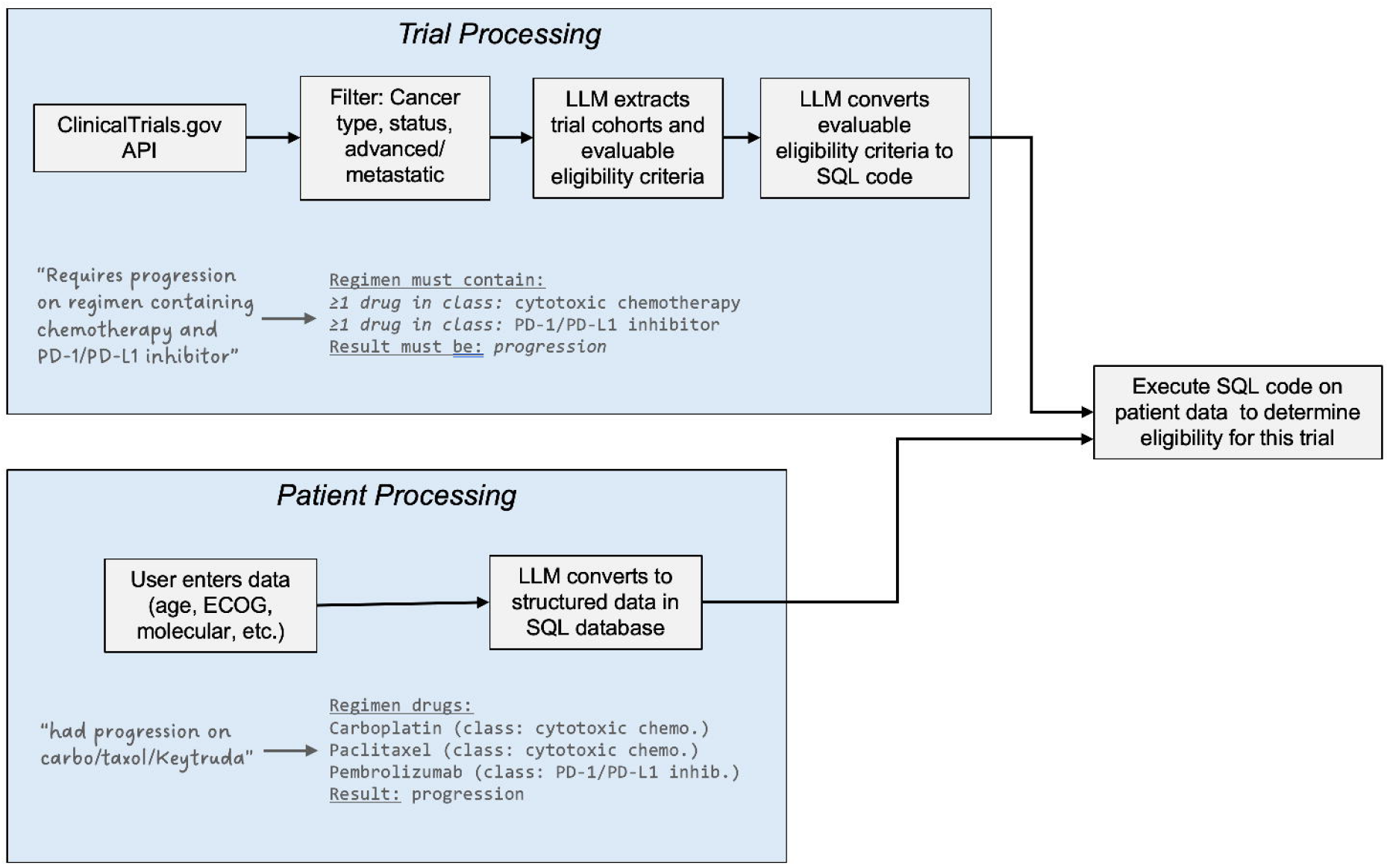
Free text to SQL trial matching pipeline overview. LLM=large language model.

Active clinical trials as of July 14, 2025 were downloaded from ClinicalTrials.gov. Studies were included if they were interventional, had the primary purpose of treatment, had at least one site in the United States, and had a status of “actively recruiting.” Trials for breast cancer, colon cancer, and NSCLC were included and identified by searching “breast cancer”, “colon cancer”, and “non-small cell lung cancer”. Some trials targeted more than one of these cancer types, such as many phase 1 trials. We excluded trials not for advanced/metastatic disease using OpenAI GPT-4.1 with a custom prompt.

### Synthetic patients

Board-certified medical oncologists who treat breast, lung, and colon cancer constructed clinically relevant synthetic patient vignettes, eight per disease group. To capture a wide variety of common scenarios, we varied molecular findings, treatment history, performance status, and age.

### Matching process

Before SQL generation, Gemini 2.5 Pro separated each trial’s eligibility criteria into evaluable and unevaluable components based on the available patient data fields and extracted trial cohort definitions. A trained research assistant reviewed all cohort definitions and evaluable/unevaluable criteria splits and edited them as needed. Trial eligibility criteria (only evaluable components) were converted into SQL using Gemini 2.5 Pro. Synthetic patient vignettes were structured through the TrialFetch chat interface using GPT-5 mini, and the resulting records were reviewed by a physician and edited as needed. Matching then executed saved SQL against these records without further LLM inference. Implementation details and generation settings are provided in the Supplementary Methods and Table S3.

We used the following variables to determine if a synthetic patient was likely eligible for a trial: age, sex, ECOG performance status, cancer type (solid tumor, lymphoma, etc.), primary site (lung, breast, etc.), histology, extent (localized, locoregional, metastatic), presence of active brain metastases, molecular findings including genes mutated (e.g., *KRAS* but not specific subtypes such as G12C vs. G12D/V) and immunohistochemistry, systemic therapy regimens in order (including drugs in each regimen, and response to each regimen, coded as progression/no progression). Eligibility criteria often require or exclude past treatment with a certain drug class such as EGFR inhibitors; to enable this we included each drug’s World Health Organization Anatomical Therapeutic Chemical (ATC) classification in its database record.^14^ Further details of eligibility determination are found in the Supplementary Methods.

For filtering based on whether there was a close trial site to the patient, a distance of 100 miles from the patient’s ZIP code was used, based on a previous study that found that most cancer patients would travel up to 100 miles for a trial.^15^

### Descriptive eligibility analysis

For a patient-level analysis, we summarized potential eligibility descriptively for the 24 synthetic patient vignettes. For the patient-level scenario analyses, we varied one characteristic at a time (ECOG performance status, age, sex, or ZIP code), held the remaining characteristics fixed, and plotted the mean number of potentially eligible trials across the same 24 vignettes. ZIP-code analyses counted only trials with a site within 100 miles of the specified ZIP code. These were illustrative location scenarios, with equal weight given to each vignette and no population weighting.

Then, for a trial-level analysis, for each trial we counted how many of the 24 vignettes were potentially eligible using their original characteristics and without geographic restriction. We summarized these counts using medians and interquartile ranges (IQRs), overall and by trial phase, design allocation, and the presence of molecular criteria. Quartiles were calculated by linear interpolation.

Analysis was performed from January through September 2026 using R version 4.5.1 and Python version 3.12.14.

## Results

### Trials

There were 833 trials for breast cancer, colon cancer, and NSCLC in ClinicalTrials.gov with interventional study type and purpose of “treatment”. Two physicians independently classified a random sample of 50 trials by disease setting (curative, advanced/metastatic, both, or uncertain), blinded to the automated classifications and each other’s ratings. Initial physician agreement was 45/50 (90.0%; Cohen’s κ=0.794). After consensus adjudication, two trials remained uncertain and were excluded from the comparison. Automated classifications agreed with consensus on 45/48 evaluable trials (93.8%; κ=0.864). See the Supplementary Methods and Supplementary Results for methods and discrepancies.

Using the LLM classifications for all 833 trials, seventy-three trials were for curative intent, and 14 were for both curative and advanced/metastatic, leaving 746 eligible trials for advanced/metastatic disease which were analyzed further. These trials most commonly targeted NSCLC (56.6%), followed by breast cancer (43.6%) and colon cancer (24.8%); each trial could have more than one designated cancer type. Most trials were in phase 1 (35.8%). Most trials were sponsored by industry (64.1%), followed by other sources (30.6%, not broken down further in ClinicalTrials.gov), then federal (5.4%). Finally, most trials were started recently (2022-present constituted 73.5%) (Table 1).

**Table 1.** Features of 746 Clinical Trials.

| Variable | Count (%) or Median (Q1, Q3) |
| --- | --- |
| Cancer |  |
| Breast cancer | 325 (43.6%)* |
| Colon cancer | 185 (24.8%)* |
| Non-small cell lung cancer | 422 (56.6%)* |
| Phase |  |
| 1 | 267 (35.8%) |
| 1/2 | 192 (25.7%) |
| 2 | 184 (24.7%) |
| 2/3 | 8 (1.1%) |
| 3 | 82 (11%) |
| 4 | 3 (0.4%) |
| Not stated | 10 (1.3%) |
| Sponsor class |  |
| Federal | 40 (5.4%) |
| Industry | 478 (64.1%) |
| Other (academic, etc.) | 228 (30.6%) |
| Design allocation |  |
| Non-randomized | 309 (41%) |
| Randomized | 196 (26%) |
| Not stated | 241 (32%) |
| Planned enrollment | 124 (50, 280) |
| Eligibility criteria include a molecular finding | 613 (82%) |
| Year started |  |
| 2010-2011 | 1 (0.13%) |
| 2012-2013 | 2 (0.26%) |
| 2014-2015 | 3 (0.40%) |
| 2016-2017 | 16 (2.1%) |
| 2018-2019 | 51 (6.9%) |
| 2020-2021 | 125 (16.8%) |
| 2022-2023 | 288 (38.6%) |
| 2024-2025 | 260 (34.9%) |

\* Percentages do not add up to 100% since one trial could target multiple cancer types

### Internal Validation of Eligibility Determination

During preprocessing, the research assistant edited 65 cohort definitions and 6 evaluable/unevaluable criteria splits.

We created 24 synthetic patients with metastatic breast, colon, or NSCLC (Table S1). To examine the generated SQL and the effect of physician edits on matching results, we had a physician review and edit the computer-generated code for 30 trials. Then, for each combination of trial and patient (720), we assessed whether the patient was eligible for the trial according to the initial computer code, and then according to the physician-edited code. The physician made edits to the code for 11 of 30 trials. Examples of such edits included changing the result in situations when the criterion is not fully evaluable with available data, such as counting adjuvant chemotherapy as a line of therapy if it was given <6 months prior to recurrence. A description of all the edits is in Table S2. Table S4 shows SQL for two trials, one approved by the physician without edits and one with physician edits shown. For 149/720 combinations (21%), the patient was eligible according to the computer code. The original and edited code produced identical classifications for all 720 pairs.

For manual eligibility validation, we randomly selected ten trials, with two trials from each quintile of automated eligible-patient count. Two physicians independently assessed all 240 patient-trial pairs using the same limited patient variables and evaluable criteria as the AI system, with disagreements resolved by consensus. Initial physician agreement was 214/240 (89.2%; κ=0.682). Physician consensus classified 60 patient-trial pairs as potentially eligible and 180 as ineligible. Automated matching agreed with consensus on 230/240 pairs (95.8%; κ=0.882), with 50 true positives, 180 true negatives, no false positives, and ten false negatives. Sensitivity was 83.3%, specificity 100%, positive predictive value 100%, and negative predictive value 94.7%. Eight of the false negatives were for a trial that required HER2 mutation or HER2 immunohistochemistry (IHC) 1-3+; for cases marked HER2 negative the LLM scored the patients as ineligible whereas the physicians scored the patients as eligible since HER2 negative can include 1+ staining. Review of the LLM prompt for eligibility determination showed that this situation was not explained clearly enough to the LLM. See the Supplementary Results for more details of discrepancies.

### Potential eligibility across patient scenarios

We examined patterns of patient eligibility for 24 synthetic patients for the 746 trials in the database. Without geographic restrictions, the number of eligible trials varied dramatically by synthetic patient, ranging from 10–258 (Figure 2, Table S1). Note that patients were often eligible for some trials that did not have a specific designation in ClinicalTrials.gov for their cancer type. This typically occurred for phase 1 trials that allowed a variety of cancer types but did not list them all in their ClinicalTrials.gov conditions list. Consequently, registry cancer designations alone did not determine which trials matched a vignette.

**Figure 2.**
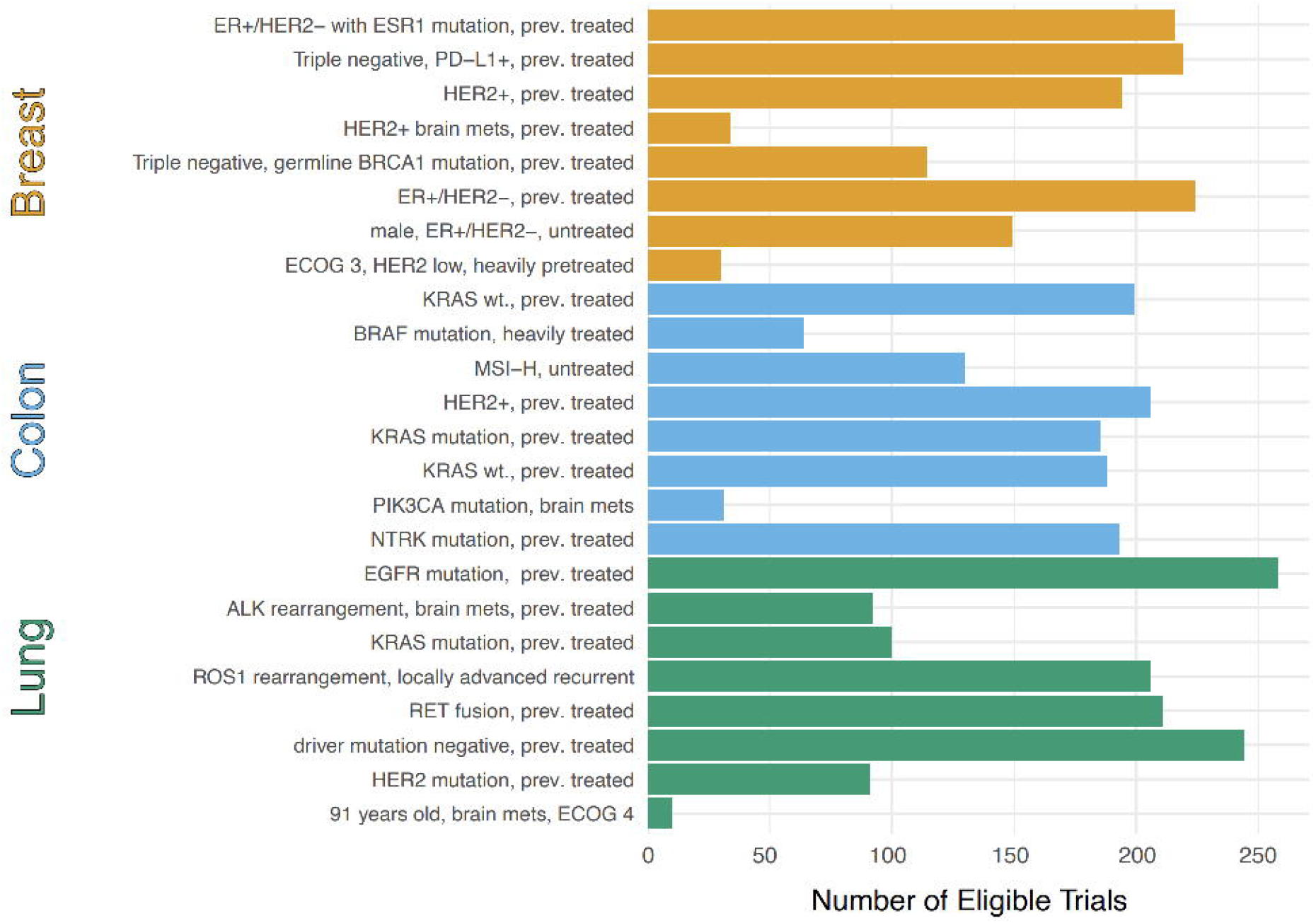
Number of eligible trials for each synthetic patient (assuming no geographic restriction), out of 746 trials for breast, colon, and non-small cell lung cancer. See Table S1 for full patient details.

We tested the impact of varying each patient variable using the following procedure: for each synthetic patient, we maintained all other variables at their existing values, varied the single variable through its range, and recorded the number of eligible trials for each value. For age, the mean number of potentially eligible trials was relatively stable across ages 30–90 (150 trials at age 30 and 146.8 at age 90); fewer trials matched at age 10 (63.2 trials) (Figure 3A). Patients with ECOG 0 qualified for an average of 187 trials, which fell sharply as ECOG worsened (ECOG 2 = 65 trials, ECOG 4 = 23 trials) (Figure 3B). Trials for male vs. female sex did not vary substantially (Figure 3C). We observed large differences by geography, when considering trials that had a site within 100 miles of different ZIP codes: for example, the New York City ZIP code yielded 80.5 trials, while the South Dakota ZIP code yielded only six (Figure 3D).

**Figure 3.**
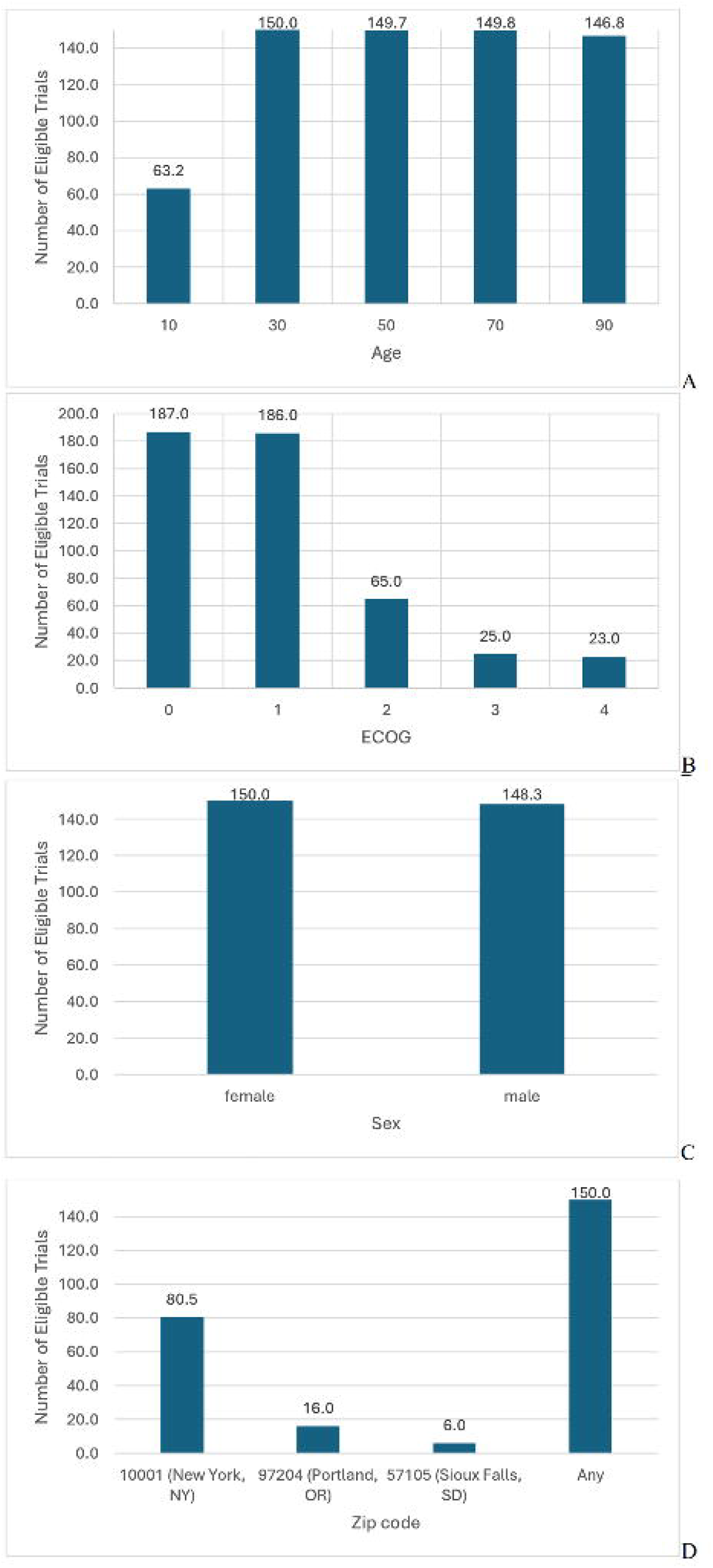
Mean number of eligible trials (out of 746) for 24 synthetic patients when varying: (A) Age; (B) ECOG performance status; (C) Sex; (D) ZIP Code. For ZIP code, we only allowed trials with a site within 100 miles of that zip code.

### Potential eligibility by trial characteristics

Across 746 trials, the median number of potentially eligible synthetic patients per trial was 2 of 24 (IQR, 0–7). Phase 1 trials had a median of 5 potentially eligible patients (IQR, 1–13), compared with 0 (IQR, 0–1) for phase 3 trials. The corresponding medians were 4 (IQR, 1–13) for nonrandomized trials and 1 (IQR, 0–2) for randomized trials, and 8 (IQR, 4–15) for trials without molecular criteria versus 1 (IQR, 0–5) for those with molecular criteria (Table 2).

**Table 2.** Potential eligibility among 24 synthetic patient vignettes, by trial characteristics.

| **Trial characteristic** | **Trials, n** | **Eligible vignettes per trial, out of 24, median (Q1–Q3)** |
| --- | --- | --- |
| All trials | 746 | 2 (0–7) |
| Phase |  |  |
| 1 | 267 | 5 (1–13) |
| 1/2 | 192 | 3 (1–11) |
| 2 | 184 | 1 (0–4) |
| 2/3 | 8 | 0 (0–0.25) |
| 3 | 82 | 0 (0–1) |
| 4 | 3 | 0 (0–0.5) |
| Not stated | 10 | 9.5 (3–18.5) |
| Design allocation |  |  |
| Nonrandomized | 309 | 4 (1–13) |
| Randomized | 196 | 1 (0–2) |
| Not stated | 241 | 2 (0–7) |
| Eligibility criteria include a molecular finding |  |  |
| No | 133 | 8 (4–15) |
| Yes | 613 | 1 (0–5) |

Q1 and Q3 denote the 25th and 75th percentiles

Per-patient eligibility determination for all trials (execution of the 746 trial SQL statements) took a median of 0.459 seconds per patient (IQR, 0.454–0.470), corresponding to 1,624 patient-trial pairs per second (IQR, 1,587–1,642). Timings excluded retrieval of trial SQL (which can be done occasionally and cached) and creation of patient SQLite records.

Details of all 746 trials, LLM prompts, and raw data for the patient- and trial-level results are available online.^16^

## Discussion

We developed a system that combines efficient trial screening with explicit eligibility logic applied to structured clinical and molecular characteristics. Automated eligibility determinations agreed with physician consensus on 95.8% of 240 patient-trial pairs spanning ten trials and 24 synthetic patients with breast, colon, and lung cancer. The key innovation is the automatic conversion of each trial’s eligibility criteria to SQL computer code using an LLM. Once a patient’s data has also been converted to structured format, which is achievable with LLMs, their eligibility for thousands of trials can be calculated in a few seconds without requiring any more LLM use. This enables cost-effective and rapid trial matching on the TrialFetch site, and also enabled the current study of many synthetic patients’ eligibility patterns. Matching against all 746 trials took 0.46 seconds per patient, which is fast enough to serve as part of a health system-wide matching system. This demonstration describes potential eligibility for 24 synthetic vignettes across a defined cohort of US trials for three cancer types identified from a single registry snapshot. Given the burden of reviewing complex, frequently changing criteria, TrialFetch uses SQL matching to identify candidates, giving patients the benefit of the doubt on partially evaluable criteria, so candidates can undergo more detailed LLM-assisted interpretation or human review of eligibility and fit with patient goals. The 83.3% sensitivity in our ten-trial evaluation shows that this screening can miss potentially eligible trials, and our results do not establish that generated SQL can safely bypass review.

There have been several prior studies of using LLMs for clinical trial eligibility determination.^8– 11^ TrialGPT retrieves candidate trials, uses an LLM to assess eligibility at the criterion level, and aggregates these assessments into trial-level rankings.^8^ PRISM uses a custom LLM (OncoLLM) to assess eligibility criteria using information retrieved from unstructured electronic health records (EHRs).^9^ TrialFetch instead executes reusable SQL against structured patient records, avoiding additional LLM inference for each patient-trial pair during this screening stage. Differences in patient inputs, evaluated criteria, and outcome definitions make it difficult to compare accuracy across these studies. Our evaluation used physician-reviewed structured synthetic records and did not test extraction from raw EHR data, an important upstream challenge.

Although EHR abstraction may itself require LLM inference, it is performed once per patient record or update and can then be reused. Similarly, each trial’s eligibility criteria are converted to SQL once and updated only when the trial changes. LLM inference therefore scales approximately with the number of patients plus trials, rather than with every patient-trial combination; subsequent many-to-many matching requires no additional LLM inference. We did not evaluate the accuracy or computational cost of the upstream EHR abstraction step.

### Patterns of eligibility

In our selected illustrative patient scenarios, the number of potentially eligible trials decreased with worse performance status and varied across the selected locations. Later-phase, randomized, and molecularly selective trials generally matched fewer vignettes. These patterns depend on vignette composition and should not be interpreted as independent effects of trial characteristics or as population estimates of trial access. The performance status finding is important because these were metastatic cancer trials, where poor functional status is common, yet many studies reserve enrollment for the fittest patients.^17,18^

The selected location scenarios also produced different matching results. Averaged across the same 24 synthetic vignettes, 6 potentially eligible trials had a site within 100 miles of the Sioux Falls, SD, ZIP code, compared with 80.5 for the New York City ZIP code. These counts do not measure actual access or enrollment opportunities. However, they are consistent with prior work showing concentration of oncology trials in urban and academic centers.^5,7^ In the future, sponsors should incorporate decentralized elements when possible, consistent with FDA guidance.^19^ For trials requiring specialized expertise or complex therapeutics, regional hub-and-spoke models could preserve oversight at high-volume centers while extending routine trial activities to community oncology practices.

These demonstrations show how the platform can summarize potential eligibility as individual inputs vary. Evaluating population-level access or policy changes would require representative patient data and information about real-world participation barriers.

### Limitations

This study had several limitations. While sponsors are required to submit trial data to ClinicalTrials.gov for all phase 2-4 drug trials in the US, not all phase 1 trials are included^20^, so some trials are likely missing from our analysis.

This was a proof-of-concept study using clinician-designed synthetic patients with metastatic breast cancer, colon cancer, and NSCLC. Although these vignettes were constructed to reflect clinically relevant scenarios, they were not intended to match the real-world distribution of clinical variables, and the findings may not generalize to other cancer types or broader patient populations. The trial-level analysis depended on the selected mix of cancer types, molecular findings, and treatment histories and could differ in a representative patient cohort. Future studies could use medical record data to generate synthetic patients whose clinical and molecular features more closely reflect real-world distributions.^21^ We restricted this initial analysis to advanced/metastatic disease because this setting has substantial unmet need, a large number of active trials, and complex eligibility requirements involving prior lines of systemic therapy and molecular findings. Earlier-stage trials often require more detailed representation of anatomic disease distribution, stage, and local therapies; supporting these variables would require expansion of the current patient schema. Extending the system to earlier-stage disease is an important direction for future work.

To simplify matching, we did not use non-cancer comorbidities, lab results, or local therapies such as surgery and radiation. In addition, when a criterion was only partially evaluable with the available structured data, the system gave the patient the benefit of the doubt on the unevaluable portion. Mutations were matched at the gene level without distinguishing specific variants, so variant-specific inclusion or exclusion criteria can yield incorrect eligibility classifications; for example, a KRAS G12D mutation could satisfy a gene-level representation of a KRAS G12C requirement. In the TrialFetch website, a subsequent LLM-assisted review can consider recorded variant-level patient details and retrieve full trial eligibility criteria to refine the initial screening results; this stage was not evaluated in the present study. Incorporating variant-specific matching directly into the screening engine is a priority for future development.

Protocol-based eligibility and geographic proximity to a trial site also do not necessarily translate into actual trial access or enrollment. Real-world participation is influenced by factors such as physician referral, site-level screening practices, trial slot availability, insurance and financial barriers, transportation, and patient preferences. Geographical proximity to the trial acts not as a strict exclusion filter but rather is factored into consideration by patients when making decisions. Our results should therefore be interpreted as estimating potential eligibility within the trial ecosystem rather than realized enrollment opportunity.

Finally, eligibility validation was limited to ten trials and 24 synthetic patient vignettes, with repeated patients and trials across the 240 pairs. Physician review identified substantive SQL edits (Table S2), but the unchanged classifications across 720 patient-trial pairs indicate limited coverage of the edited logic by these vignettes. This internal-consistency evaluation does not establish real-world clinical eligibility or validate performance on actual patient records.

In conclusion, LLM-based conversion of eligibility criteria to structured queries enables efficient screening using explicit eligibility logic, and analysis of potential clinical trial eligibility patterns. This methodology could support future research on trial access disparities, inform real-time trial matching workflows, and help evaluate how changes in eligibility criteria may affect which patients can participate in clinical trials.

## Data Availability

All data produced in the present study are available upon reasonable request to the authors.

https://doi.org/10.17605/OSF.IO/2EGSZ

## Acknowledgements

N/A

