## Supplement for "Quantifying Cancer Clinical Trial Eligibility Using Artificial Intelligence-Based Matching"

**Supplementary Material**

**Supplementary Methods**

*Trial processing*

Trial data were downloaded from ClinicalTrials.gov on July 14, 2025, and processed between that date and the beginning of the main analysis in January 2026. After filtering by cancer type, recruitment status, study type, treatment purpose, and US site availability as described in the main Methods, GPT-4.1 classified trial setting from the title and eligibility text as curative, advanced/metastatic, both, or uncertain. Trials classified as advanced/metastatic proceeded to eligibility processing. Table S3 summarizes the LLM stages, inputs, outputs, and generation settings.

Disease setting validation

For disease setting validation, two physicians independently classified a random sample of 50 trials from their titles and eligibility text. The four categories were curative (primary or locoregional disease treated with curative intent), advanced/metastatic (distant metastases or incurable locally advanced/recurrent disease), both settings, and uncertain.

We calculated initial physician–physician agreement and Cohen’s κ before adjudication. Disagreements were resolved by physician consensus using the source material. Trial-setting consensus remained uncertain in two cases; these were excluded from all consensus-versus-model metrics, with the excluded number reported. We compared automated predictions with the final reference using exact agreement and κ, and calculated sensitivity, specificity, positive predictive value, and negative predictive value for binary eligibility.

Evaluable criteria and cohort extraction

Gemini 2.5 Pro used each trial’s eligibility text and the patient data schema described below to separate criteria into evaluable and unevaluable components. Criteria based on fields represented in the schema could be evaluated computationally; requirements based entirely on unavailable fields were excluded from automated evaluation.

When a criterion was only partially evaluable with the selected data elements (e.g., "at least 12 months of clinical response to first-line systemic therapy," where we capture the regimen’s final response status but not duration of response), the system evaluated the portion it could and gave the patient the benefit of the doubt on the remainder. In the TrialFetch web site, in this situation a message is displayed prompting the user to verify the unevaluable component. The system assumes that next-generation sequencing (NGS) was performed and all cancer-relevant gene mutations are recorded; an unrecorded required mutation therefore causes the patient to be ineligible. Mutations are matched at the gene level, without requiring the specific exon or variant. This simplification can produce incorrect classifications when inclusion or exclusion depends on a particular variant; for example, a KRAS G12D mutation may satisfy a gene-level representation of a KRAS G12C requirement. Unreported non-mutation biomarkers, such as immunohistochemistry or MSI/MMR status, receive the benefit of the doubt on eligibility (though confirmatory testing would be required before clinical eligibility can be established, and a note about this is displayed on the TrialFetch site). Criteria that relied only on variables not represented in the patient schema, such as laboratory values, organ function requirements, or washout intervals were not evaluated.

Gemini 2.5 Pro then used the trial title, descriptions, arms, outcomes, and evaluable criteria to extract a list of trial cohorts and associated notes. A research assistant reviewed all cohort definitions and evaluable/unevaluable criteria splits and edited them as needed before SQL generation. Outputs from both extraction stages were parsed, with one regeneration attempt if an output was invalid.

SQL generation and selection

Using the reviewed evaluable criteria and cohort definitions, patient schema, and drug-class information, Gemini 2.5 Pro generated three candidate SQL SELECT statements in separate requests. Candidate SQL was executed against sample patient databases to detect execution errors and check the expected output format, with regeneration of invalid candidates. An additional LLM call selected one candidate. The selected statement was saved for subsequent patient matching.

*Patient data structuring*

Synthetic patient vignettes were entered through the TrialFetch chat interface and structured using GPT-5 mini with reasoning effort set to minimal. The agent used drug-name and histology lookup tools and saved structured YAML through a tool call; application code converted the YAML into database records for subsequent SQLite export. A physician reviewed the structured records in the TrialFetch interface and edited them as needed.

Patient data schema

The structured patient record comprises five linked entities. ZIP code is stored with the patient and used separately for geographic filtering.

**Patient and geographic data:** age, sex, ECOG performance status (0-4), and ZIP code.

**Cancer:** a unique identifier; an indicator for whether the cancer is used for trial matching; cancer type; primary site; ICD-O-3 histology; current extent (localized, locoregional, metastatic, or no evidence of disease); presence of active/untreated brain metastases; and years since active disease, when applicable.

**Systemic regimen:** the linked cancer; regimen order; treatment setting (neoadjuvant, adjuvant, concurrent with radiation, advanced/recurrent, or metastatic); and regimen result (progression, no progression, or unknown).

**Drug:** the linked regimen; standardized international nonproprietary drug name; and third- and fourth-level World Health Organization Anatomical Therapeutic Chemical codes.

**Molecular finding:** the linked cancer; marker name; finding type (immunohistochemistry, genomic instability, mutation/deletion, amplification, fusion/rearrangement, methylation, aneuploidy, translocation, germline mutation/deletion, or other); and result. The current schema represents mutations at the gene level rather than the specific variant level.

**Definition of regimen result:** the final disease status within 30 days after the last dose of a regimen, coded as progression, no progression, or unknown. It is not the best response achieved during the regimen.

Example structured synthetic patient

The following example illustrates the information available to the matching system for one synthetic breast cancer vignette.

**Patient:** 58-year-old woman; ECOG performance status 0; ZIP code not provided.

**Cancer:** active metastatic solid tumor of breast primary site; adenocarcinoma histology; used for trial matching; no active/untreated brain metastases.

**Molecular findings:** ER positive, PR negative, and HER2 positive by immunohistochemistry.

**Systemic therapy regimens, in order:** (1) paclitaxel, trastuzumab, pertuzumab, and anastrozole; (2) trastuzumab emtansine; (3) capecitabine, tucatinib, and trastuzumab; and (4) trastuzumab deruxtecan. Each regimen was administered in the metastatic setting and had a final result of progression.

*Eligibility matching and validation*

Each patient’s structured data were exported to an SQLite database. For each patient-trial pair, the saved SQL SELECT statement was executed against that database and returned the trial cohorts for which the patient was potentially eligible. We examined physician edits to the eligibility code and separately evaluated agreement with physician eligibility assessments:

1. For 30 trials (selected to have a mix of phases and target cancer types), a physician reviewed the computer-generated SQL code for the eligibility criteria, compared it to the eligibility criteria text, and made edits as needed. For each of the 24 synthetic patients, for each of these 30 trials we assessed whether the patient was eligible for the trial according to the initial computer code, and then according to the physician-edited code. Agreement between these versions measures the effect of the edits within the tested vignettes; it does not establish that the edits were unnecessary or that generated SQL can safely bypass review.
2. We selected ten trials by stratified random sampling across quintiles of automated eligible-patient count, yielding 240 patient-trial pairs. Two physicians rated eligibility for each of the 24 synthetic vignettes for each trial. Raters applied the same gene-level mutation matching and benefit-of-the-doubt rules for missing or partially evaluable information. Agreement with these ratings therefore does not validate variant-specific eligibility. Disease was assumed metastatic and to have progressed on each listed regimen unless the vignette stated otherwise; active brain metastases were assumed absent unless stated. Independent ratings were blinded to stored automated predictions and the other physician’s ratings.

*Eligibility execution benchmark*

We benchmarked execution of the pre-generated trial eligibility SQL statements for the 746-trial study cohort. The trial statements were retrieved from the application’s PostgreSQL database before timing. Each synthetic patient’s structured data were exported to a local SQLite database before timing. For each patient, all 746 SQL statements were executed sequentially using a single SQLite connection; this process was repeated three times. The benchmark did not include retrieval of trial SQL statements, patient-data export, cache operations, task orchestration, or storage of matching results. It was performed on a MacBook Pro with an Apple M1 Pro processor (8 cores), 32 GB memory, Python 3.12.12, and SQLite 3.50.4. For each patient, we calculated the median runtime across the three repetitions and then summarized these patient-level medians across the 24 patients using the median and interquartile range.

*LLM implementation and reproducibility*

Generation parameters were left at provider defaults unless specified in Table S3; no random seed was supplied. For the Gemini 2.5 Pro model used to generate the SQL code, parameters were: temperature 1.0, top-p 0.95, and fixed top-k 64.

Execution of fixed SQL against fixed structured patient records requires no additional LLM inference. Repeating the upstream LLM generation could produce different outputs; identical regenerated SQL was not established. Reproduction of matching results requires the same SQL statements, patient records, schema, reference data, and execution environment.

*Search sensitivity analysis*

Using only one ClinicalTrials.gov search term for each cancer type to retrieve trials, as we did for this study, could potentially miss some relevant trials. To investigate this, on September 17, 2026, we assessed the sensitivity of trial retrieval to the search terms "colon cancer" (used for the study) and "colorectal cancer" (not used for the study) using the ClinicalTrials.gov API condition-search field. We used the same filters as used in our study: recruiting status, interventional study type, treatment as the primary purpose, and at least one US site. The condition-search field incorporates synonym and MeSH-related indexing; we compared the results returned by each term and their deduplicated unions. For additional trials not returned by "colon cancer", we reviewed their descriptions and eligibility criteria for inclusion of colon cancer and advanced/metastatic setting. We restricted these additional trials to those first posted on or before July 14, 2025 (the date of the main ClinicalTrials.gov data pull for this study).

**Supplementary Results**

*Search sensitivity analysis*

The "colon cancer" search returned 244 trials. Adding "colorectal cancer" increased the union to 306 trials, with 62 additional trials. Of the 62 additional trials, 41 were first posted on or before July 14, 2025. Seventeen met the colon-cancer and disease-setting criteria; 14 were already represented in the 746-trial cohort. The three remaining trials were NCT05719558 (ASP1002), NCT05947838 (colorectal peritoneal metastases), and NCT06934538 (STC-1010/BreAK CRC). The findings indicate that using "colon cancer" alone may miss a few relevant trials registered under broader colorectal terminology. Because this check used contemporary registry records, these are potential rather than confirmed omissions from the July 2025 snapshot.

*Physician validation of early stage vs. metastatic setting*

The two physicians initially agreed on 45/50 disease-setting classifications (90.0%; κ=0.794). Following consensus, 32 trials were advanced/metastatic, 12 curative, four both settings, and two uncertain. The uncertain trials were NCT06046495 (PLB1004 in NSCLC) and NCT06413680 (REGN10597 in solid tumors), for which the available descriptions did not permit a definitive consensus classification. Excluding these two trials, exact automated–consensus agreement was 45/48 (93.8%; κ=0.864).

All three discrepancies had consensus “both curative and advanced/metastatic.” For NCT04789486 (Nano-SMART), the automated classification was curative; the trial includes node-negative NSCLC and locally advanced unresectable pancreatic cancer, which the physicians interpreted as spanning both settings. For NCT04821284 (ultrasound-enhanced drug delivery in pancreatic cancer), the automated classification was advanced/metastatic; the criteria include stage II/III and stage IV disease, which the physicians classified as both settings, although the condensed eligibility text does not explicitly define resectability. For NCT01804686 (ibrutinib long-term extension), the automated classification was advanced/metastatic and consensus was both settings. Its eligibility depends on participation in prior ibrutinib trials, most of which are for incurable disease, rather than an explicit stage or treatment-intent restriction. These cases illustrate ambiguity in assigning a single setting to heterogeneous trial populations.

*Physician validation of eligibility*

The physicians initially agreed on 214/240 eligibility decisions (89.2%; κ=0.682), with 26 disagreements resolved by consensus. Consensus classified 60 pairs as potentially eligible and 180 as ineligible. Against this reference, the stored automated predictions yielded 50 true positives, 180 true negatives, no false positives, and ten false negatives: agreement 230/240 (95.8%), sensitivity 50/60 (83.3%), specificity 180/180 (100%), positive predictive value 50/50 (100%), negative predictive value 180/190 (94.7%), and κ=0.882. Eight trials had complete agreement across all 24 patients; NCT06714617 had 16/24 agreement and NCT05838768 had 22/24.

Eight false negatives occurred in NCT06714617 (BL-M17D1), which accepts HER2 expression at IHC 1+ to 3+ or qualifying HER2 alterations. All eight vignettes recorded HER2-negative disease without an IHC score. HER2 1+ is ambiguous since in a binary classification it is negative, but for this trial it was considered positive. The generated SQL treated a negative HER2 IHC value as ineligible, whereas the physicians considered the trial-specific expression threshold unresolved and applied the benefit-of-the-doubt rule: a general HER2-negative designation does not distinguish IHC 0 from low expression meeting this trial’s threshold.

The remaining two false negatives occurred in NCT05838768 (HRO761). In nsclc_test_pt_5, repotrectinib had no recorded drug-class code, so the SQL did not recognize the prior targeted therapy required for Arm B. In bc_test_pt_7, the broad cancer-type field was “breast” rather than “solid_tumor,” causing failure of the solid-tumor condition despite the correct breast primary site.

**Supplementary Tables**

Table S1: Trial Eligibility of Synthetic Patients With Breast, Colon, or Non-Small Cell Lung Cancer

| **Primary site** | **Histology** | **Age** | **Sex** | **ECOG PS** | **Molecular findings** | **Prior Treatment(s)^*^** | **Misc.** | **Num. eligible trials** |
| --- | --- | --- | --- | --- | --- | --- | --- | --- |
| Breast | adenocarcinoma | 72 | F | 1 | ER+; PR+; HER2-; ESR1 mutation | Anastrozole/Ribociclib; Elacestrant | Localized stomach cancer 3 years ago, NED | 216 |
|  | adenocarcinoma | 45 | F | 1 | ER-; PR-; HER2-; PD-L1 15% | Carboplatin/Gemcitabine/Pembrolizumab; Sacituzumab Govitecan |  | 219 |
|  | adenocarcinoma | 58 | F | 0 | ER+; PR-; HER2+ | Paclitaxel/Trastuzumab/Pertuzumab/anastrozole; Trastuzumab emtansine (T-DM1); Capecitabine/tucatinib/trastuzumab; Trastuzumab deruxtecan (T-DXd) |  | 194 |
|  | adenocarcinoma | 61 | F | 2 | ER-; PR-; HER2+ | Docetaxel/Carboplatin/Trastuzumab/Pertuzumab; Trastuzumab deruxtecan (T-DXd) | Active brain mets | 34 |
|  | adenocarcinoma | 39 | F | 1 | ER-; PR-; HER2-; BRCA1 germline mutation; PD-L1 <1% | Carboplatin/Paclitaxel; Olaparib |  | 114 |
|  | adenocarcinoma | 65 | F | 1 | ER+; PR+; HER2-; No actionable mutations found | Letrozole/Ibrance; Fulvestrant/Everolimus |  | 224 |
|  | adenocarcinoma | 75 | M | 1 | ER+; PR+; HER2-; BRCA2 germline mutation | none |  | 149 |
|  | adenocarcinoma | 53 | F | 3 | ER+; PR-; HER2-; AKT1 mutation | Letrozole/Palbociclib; Fulvestrant/Capivasertib; Capecitabine; Trastuzumab deruxtecan (T-DXd) |  | 30 |
| Colon | adenocarcinoma | 58 | M | 1 | KRAS/NRAS/BRAF wt (left-sided); MSS; HER2- | FOLFOX + panitumumab; FOLFIRI + bevacizumab |  | 199 |
|  | adenocarcinoma | 71 | F | 2 | BRAF V600E; MSS; HER2- | FOLFOX/encorafenib/cetuximab; FOLFIRI |  | 64 |
|  | adenocarcinoma | 62 | M | 1 | MSI-H; NRAS mutated; BRAF wt; HER2- | none |  | 130 |
|  | adenocarcinoma | 55 | F | 1 | KRAS/NRAS/BRAF wt (right-sided); MSS; HER2+ (IHC 3+) | FOLFOX; tucatinib+trastuzumab |  | 206 |
|  | adenocarcinoma | 67 | M | 1 | KRAS G12V; MSS; BRAF wt; HER2- | FOLFIRI + bevacizumab; FOLFOX | Also has metastatic prostate cancer | 185 |
|  | adenocarcinoma | 74 | M | 0 | KRAS/NRAS/BRAF wt (right-sided); MSS; HER2- | FOLFOX + bevacizumab |  | 188 |
|  | adenocarcinoma | 60 | F | 2 | KRAS G13D; PIK3CA H1047R; MSS; HER2- | FOLFIRI + bevacizumab; FOLFOX | Active brain mets | 31 |
|  | adenocarcinoma | 45 | M | 1 | NTRK1 fusion; MSS; KRAS/NRAS/BRAF wt; HER2- | FOLFOX; FOLFIRI |  | 193 |
| Lung | adenocarcinoma | 63 | F | 1 | EGFR mutation; PD-L1 5% | Osimertinib; Carboplatin/pemetrexed/bevacizumab |  | 258 |
|  | adenocarcinoma | 54 | M | 0 | ALK fusion; PD-L1 70% | alectinib; lorlatinib | Active brain mets | 92 |
|  | adenocarcinoma | 71 | F | 2 | KRAS mutation; STK11 mutation; PD-L1 80% | pembrolizumab/carboplatin/pemetrexed; adagrasib without progression | Had localized breast cancer 4 years ago treated with surgery alone with NED | 100 |
|  | adenocarcinoma | 86 | F | 0 | ROS1 fusion; PD-L1 <1% | repotrectinib for recurrent locally advanced disease |  | 206 |
|  | adenocarcinoma | 65 | M | 1 | RET fusion; PD-L1 30% | selpercatinib; carbo/taxol/pembrolizumab | Untreated prostate cancer | 211 |
|  | squamous cell carcinoma | 69 | F | 1 | PD-L1 90%; TMB high | pembrolizumab | Had ovarian cancer 6 years ago with NED | 244 |
|  | adenocarcinoma | 46 | M | 2 | HER2 mutation; PD-L1 5% | trastuzumab deruxtecan; carboplatin/pemetrexed/bevacizumab/atezolizumab |  | 91 |
|  | large cell neuroendocrine carcinoma | 91 | M | 4 | none | none | Has active brain mets | 10 |

PS=performance status. NED=no evidence of disease. IHC=immunohistochemistry.

* Assume for metastatic disease and had progression unless otherwise stated. Regimens separated by semicolon.

Table S2: Physician edits to computer code for eligibility determination for 30 trials. Edits were made for 11 of 30 trials.

| **NCT ID** | **Change** |
| --- | --- |
| NCT06625775 | For HER2+ breast cancer, don't exclude for more than 1 prior line of any monoclonal antibody, only if more than 1 line of ADCs specifically. There is no ATC code for ADCs specifically, so use the ADC drug names commonly used for breast cancer: trastuzumab emtansine, trastuzumab deruxtecan, datopotamab deruxtecan, sacituzumab govitecan |
| NCT06907615 | For this trial, the eligibility criteria were left blank in the prompt for the LLM due to an ingestion error, leading to the LLM hallucinating eligibility criteria. The error has been fixed, and the remaining results in the paper were run after the fix. |
| NCT06875310 | Administrative change to edit root cohort name to standard name |
| NCT06495125 | The patient must have had progression on both the immunotherapy line of treatment and the chemotherapy line of treatment. |
| NCT06881784 | Administrative change to edit root cohort name to standard name |
| NCT04429087 | Other active malignancy may not interfere with the prognosis/treatment. So allow it, but if present, add to required details to manually check, as would need to confirm would not interfere with prognosis/treatment of study cancer |
| NCT05633602 | Progression on or after platinum-based chemotherapy is not evaluable using the structured data, can only evaluate progression that happened while on the regimen so do not require progression while on platinum-based chemotherapy |
| NCT02133196 | For cervical cancer cohort, modified the required_details text about things to manually check so it is clear that if there is more than one item present the patient only needs to meet one of them. |
| NCT06750484 | Added the HER2-targeted ADCs trastuzumab emtansine and trastuzumab deruxtecan to metastatic_lines and had_prior_anti_her2 counts. Don't disqualify the patient if has_adjuvant_chemo=1 and metastatic_lines=2, because recurrence may have occurred >6 months after the last dose of (neo)adjuvant chemo. |
| NCT05142189 | Cohort 1 does not need TPS >=1% since that only applies to patients who are to start cemiplimab at cycle 3. Cohort 2 don't require progression on anti-PD-1 therapy since the progression can be up to 6 months after the treatment, and our structured data only records if there was progression while receiving the therapy. |
| NCT04585750 | if 12-17 years old, required details to manually verify should include that Safety Review Committee approval is required for ages 12-17 |

Table S3. LLM implementation and generation settings.

| **Stage** | **Model identifier** | **Input and output** | **Settings and checks** |
| --- | --- | --- | --- |
| Trial-setting classification | gpt-4.1 | Trial title and eligibility text → curative, advanced/metastatic, both, or uncertain | Provider defaults; structured response constrained to the classification categories |
| Evaluable-criteria extraction | gemini-2.5-pro | Trial eligibility text and patient schema → evaluable and nonevaluable criteria | Provider defaults; response parsing, with one regeneration attempt if batch output was invalid; research assistant review and editing of all outputs before SQL generation |
| Cohort extraction | gemini-2.5-pro | Trial title, descriptions, arms, outcomes, and evaluable criteria → cohort list and notes | Provider defaults; structured-output parsing, with one regeneration attempt if batch output was invalid; research assistant review and editing of all outputs before SQL generation |
| SQL generation and selection | gemini-2.5-pro | Evaluable criteria, cohorts, patient schema, and drug-class information → three candidates and one selected SQL statement | Provider defaults; three separate candidate requests. Execution/output checks; up to three regeneration attempts per invalid batch candidate; one retry for an invalid selection response |
| Patient structuring | gpt-5-mini | Vignette and chat exchanges, patient schema, and lookup tools → YAML saved as structured patient records | Reasoning effort: minimal. Temperature not explicitly set. Physician review and editing as needed |

**Table S4. Trial eligibility SQL before and after physician review.**

Examples from the 30-trial physician review set: (A) SQL retained without edits; (B) physician edits to a second trial. Deleted SQL is shown with strikethrough, added SQL in red, and unchanged SQL in black.

For NCT05142189, physician edits removed the general PD-L1 TPS ≥1% requirement from cohorts 1A/1B (applicable only when starting cemiplimab at cycle 3) and the cohort 2 requirement for recorded progression on immunotherapy, because qualifying progression could occur after treatment. Each statement runs against an individual patient’s SQLite database.

**A. NCT05853575 — Reviewed without edits**

A Randomized Study of Two Dosing Regimens of Adagrasib in Patients With Previously Treated Non-Small Cell Lung Cancer With KRAS G12C Mutation

```
SELECT
  'root_cohort' AS cohort_name,
  'Requires KRAS G12C mutation' AS required_details
FROM
  patient p,
  cancer c
WHERE
  c.use_for_trial_matching = TRUE
  AND p.age >= 18
  -- Inclusion: Advanced or metastatic NSCLC
  AND c.primary_site = 'lung'
  AND c.current_extent IN ('locoregional', 'metastatic')
  AND c.histology IS NOT NULL AND LOWER(c.histology) NOT LIKE 'small cell%'
  -- Inclusion: KRAS G12C mutation is required. Check for a general KRAS mutation; if positive, flag the specific variant requirement.
  AND EXISTS (
    SELECT 1
    FROM molecular_finding mf
    WHERE mf.cancer = c.id
      AND mf.name = 'KRAS'
      AND mf.finding_type = 'mutation_or_deletion'
      AND mf.result = 'positive'
  )
  -- Inclusion: Prior platinum chemotherapy
  AND EXISTS (
    SELECT 1
    FROM systemic_regimen sr
    JOIN systemic_drug sd ON sr.id = sd.regimen
    WHERE sr.cancer = c.id
      AND sd.atc_fourth_level_code = 'L01XA'
  )
  -- Inclusion: Prior immune checkpoint inhibitor
  AND EXISTS (
    SELECT 1
    FROM systemic_regimen sr
    JOIN systemic_drug sd ON sr.id = sd.regimen
    WHERE sr.cancer = c.id
      AND (sd.atc_fourth_level_code = 'L01FF' OR sd.drug_name IN ('ipilimumab', 'tremelimumab'))
  )
  -- Exclusion: Prior treatment with a KRAS G12C targeting drug
  AND NOT EXISTS (
    SELECT 1
    FROM systemic_regimen sr
    JOIN systemic_drug sd ON sr.id = sd.regimen
    WHERE sr.cancer = c.id
      AND sd.drug_name IN ('sotorasib', 'adagrasib')
  )
```

**B. NCT05142189 — SQL with physician edits shown**

LuCa-MERIT-1: First-in-human, Open Label, Phase I Dose Confirmation Trial Evaluating the Safety, Tolerability and Preliminary Efficacy of BNT116 Alone and in Combinations in Patients With Advanced Non-small Cell Lung Cancer

```
WITH PatientMainCancer AS (
    SELECT
        p.age,
        p.sex,
        p.ecog_ps,
        c.id AS cancer_id,
        c.cancer_type,
        c.primary_site,
        c.histology,
        c.current_extent,
        c.has_active_brain_mets,
        c.years_since_active_disease
    FROM patient p, cancer c
    WHERE c.use_for_trial_matching = TRUE
)
SELECT
    'Cohort 1A' AS cohort_name,
    CASE
        WHEN NOT EXISTS (SELECT 1 FROM molecular_finding mf WHERE mf.cancer = (SELECT cancer_id FROM PatientMainCancer) AND mf.name = 'PD-L1')
        THEN 'Requires PD-L1 TPS >=1%'
        ELSE NULL
    END AS required_details
    'Cohort 1A' AS cohort_name, NULL AS required_details
FROM PatientMainCancer pmc
WHERE
    -- General NSCLC
    pmc.primary_site = 'lung' AND (pmc.histology IS NOT NULL AND LOWER(pmc.histology) NOT LIKE 'small cell%')
    -- General Driver Mutation Exclusion
    AND NOT EXISTS (
        SELECT 1 FROM molecular_finding mf_drv
        WHERE mf_drv.cancer = pmc.cancer_id
        AND mf_drv.name IN ('EGFR', 'ALK', 'ROS1', 'BRAF', 'KRAS', 'MET', 'RET', 'HER2', 'NTRK1', 'NTRK2', 'NTRK3')
        AND mf_drv.finding_type IN ('mutation_or_deletion', 'fusion_or_rearrangement')
        AND mf_drv.result = 'positive'
    )
    -- General Brain Mets Exclusion (has_active_brain_mets means UNTREATED, so must be FALSE or NULL)
    AND (pmc.has_active_brain_mets IS FALSE)

    -- Cohort 1 Specific: Stage (Unresectable Stage III or metastatic Stage IV)
    AND pmc.current_extent IN ('locoregional', 'metastatic')
    -- Cohort 1 Specific: ECOG
    AND pmc.ecog_ps <= 2
    -- Cohort 1 Specific: Measurable disease NOT required.
    -- Cohort 1 Specific: Prior therapy (PD-1/L1 and Platinum)
    AND EXISTS (SELECT 1 FROM systemic_drug sd JOIN systemic_regimen sr ON sd.regimen = sr.id WHERE sr.cancer = pmc.cancer_id AND sd.atc_fourth_level_code = 'L01FF')
    AND EXISTS (SELECT 1 FROM systemic_drug sd JOIN systemic_regimen sr ON sd.regimen = sr.id WHERE sr.cancer = pmc.cancer_id AND sd.atc_fourth_level_code = 'L01XA')
    -- Cohort 1 Specific: PD-L1 TPS >=1% for those starting cemiplimab (applied as general cohort entry criterion for simplicity here)
    -- Ineligible if PD-L1 known and < 1%
    AND (
        NOT EXISTS (SELECT 1 FROM molecular_finding mf_pdl1 WHERE mf_pdl1.cancer = pmc.cancer_id AND mf_pdl1.name = 'PD-L1' AND CAST(mf_pdl1.result AS INTEGER) < 1)
    )

UNION ALL

SELECT
    'Cohort 1B' AS cohort_name,
    CASE
        WHEN NOT EXISTS (SELECT 1 FROM molecular_finding mf WHERE mf.cancer = (SELECT cancer_id FROM PatientMainCancer) AND mf.name = 'PD-L1')
        THEN 'Requires PD-L1 TPS >=1%'
        ELSE NULL
    END AS required_details
    'Cohort 1B' AS cohort_name, NULL AS required_details
FROM PatientMainCancer pmc
WHERE
    -- General NSCLC
    pmc.primary_site = 'lung' AND (pmc.histology IS NOT NULL AND LOWER(pmc.histology) NOT LIKE 'small cell%')
    -- General Driver Mutation Exclusion
    AND NOT EXISTS (
        SELECT 1 FROM molecular_finding mf_drv
        WHERE mf_drv.cancer = pmc.cancer_id
        AND mf_drv.name IN ('EGFR', 'ALK', 'ROS1', 'BRAF', 'KRAS', 'MET', 'RET', 'HER2', 'NTRK1', 'NTRK2', 'NTRK3')
        AND mf_drv.finding_type IN ('mutation_or_deletion', 'fusion_or_rearrangement')
        AND mf_drv.result = 'positive'
    )
    -- General Brain Mets Exclusion
    AND (pmc.has_active_brain_mets IS FALSE)

    -- Cohort 1 Specific: Stage
    AND pmc.current_extent IN ('locoregional', 'metastatic')
    -- Cohort 1 Specific: ECOG
    AND pmc.ecog_ps <= 2
    -- Cohort 1 Specific: Prior therapy (PD-1/L1 and Platinum)
    AND EXISTS (SELECT 1 FROM systemic_drug sd JOIN systemic_regimen sr ON sd.regimen = sr.id WHERE sr.cancer = pmc.cancer_id AND sd.atc_fourth_level_code = 'L01FF')
    AND EXISTS (SELECT 1 FROM systemic_drug sd JOIN systemic_regimen sr ON sd.regimen = sr.id WHERE sr.cancer = pmc.cancer_id AND sd.atc_fourth_level_code = 'L01XA')
    -- Cohort 1 Specific: PD-L1 TPS >=1%
    AND (
        NOT EXISTS (SELECT 1 FROM molecular_finding mf_pdl1 WHERE mf_pdl1.cancer = pmc.cancer_id AND mf_pdl1.name = 'PD-L1' AND CAST(mf_pdl1.result AS INTEGER) < 1)
    )

UNION ALL

SELECT
    'Cohort 2' AS cohort_name,
    CASE
        WHEN NOT EXISTS (SELECT 1 FROM molecular_finding mf WHERE mf.cancer = (SELECT cancer_id FROM PatientMainCancer) AND mf.name = 'PD-L1')
        THEN 'Requires PD-L1 TPS >=50%'
        ELSE NULL
    END AS required_details
FROM PatientMainCancer pmc
WHERE
    -- General NSCLC
    pmc.primary_site = 'lung' AND (pmc.histology IS NOT NULL AND LOWER(pmc.histology) NOT LIKE 'small cell%')
    -- General Driver Mutation Exclusion
    AND NOT EXISTS (
        SELECT 1 FROM molecular_finding mf_drv
        WHERE mf_drv.cancer = pmc.cancer_id
        AND mf_drv.name IN ('EGFR', 'ALK', 'ROS1', 'BRAF', 'KRAS', 'MET', 'RET', 'HER2', 'NTRK1', 'NTRK2', 'NTRK3')
        AND mf_drv.finding_type IN ('mutation_or_deletion', 'fusion_or_rearrangement')
        AND mf_drv.result = 'positive'
    )
    -- General Brain Mets Exclusion
    AND (pmc.has_active_brain_mets IS FALSE)

    -- General Cohort Criteria: Stage (Unresectable Stage III or metastatic Stage IV)
    AND pmc.current_extent IN ('locoregional', 'metastatic')
    -- General Cohort Criteria: ECOG
    AND pmc.ecog_ps <= 1
    -- Cohort Specific: Measurable Disease
    AND pmc.current_extent != 'no_evidence_of_disease'
    -- Cohort Specific: PD-L1 TPS >=50%
    AND (
        NOT EXISTS (SELECT 1 FROM molecular_finding mf_pdl1 WHERE mf_pdl1.cancer = pmc.cancer_id AND mf_pdl1.name = 'PD-L1' AND CAST(mf_pdl1.result AS INTEGER) < 50)
    )
    -- Cohort Specific: Progressive disease on/after PD-1/PD-L1 inhibitor therapy
    AND EXISTS (
        SELECT 1 FROM systemic_regimen sr
        JOIN systemic_drug sd ON sr.id = sd.regimen
        WHERE sr.cancer = pmc.cancer_id AND sd.atc_fourth_level_code = 'L01FF' AND sr.result = 'progression'
        WHERE sr.cancer = pmc.cancer_id AND sd.atc_fourth_level_code = 'L01FF'
    )

UNION ALL

SELECT
    'Cohort 3' AS cohort_name,
    NULL AS required_details -- No specific molecular markers required by name for Cohort 3 beyond general exclusions
FROM PatientMainCancer pmc
WHERE
    -- General NSCLC
    pmc.primary_site = 'lung' AND (pmc.histology IS NOT NULL AND LOWER(pmc.histology) NOT LIKE 'small cell%')
    -- General Driver Mutation Exclusion
    AND NOT EXISTS (
        SELECT 1 FROM molecular_finding mf_drv
        WHERE mf_drv.cancer = pmc.cancer_id
        AND mf_drv.name IN ('EGFR', 'ALK', 'ROS1', 'BRAF', 'KRAS', 'MET', 'RET', 'HER2', 'NTRK1', 'NTRK2', 'NTRK3')
        AND mf_drv.finding_type IN ('mutation_or_deletion', 'fusion_or_rearrangement')
        AND mf_drv.result = 'positive'
    )
    -- General Brain Mets Exclusion
    AND (pmc.has_active_brain_mets IS FALSE)

    -- General Cohort Criteria: Stage
    AND pmc.current_extent IN ('locoregional', 'metastatic')
    -- General Cohort Criteria: ECOG
    AND pmc.ecog_ps <= 1
    -- Cohort Specific: Measurable Disease
    AND pmc.current_extent != 'no_evidence_of_disease'
    -- Cohort Specific: Prior therapy (PD-1/L1 and Platinum)
    AND EXISTS (SELECT 1 FROM systemic_drug sd JOIN systemic_regimen sr ON sd.regimen = sr.id WHERE sr.cancer = pmc.cancer_id AND sd.atc_fourth_level_code = 'L01FF')
    AND EXISTS (SELECT 1 FROM systemic_drug sd JOIN systemic_regimen sr ON sd.regimen = sr.id WHERE sr.cancer = pmc.cancer_id AND sd.atc_fourth_level_code = 'L01XA')
    -- Cohort Specific: Must present with progressive disease
    AND EXISTS (SELECT 1 FROM systemic_regimen sr WHERE sr.cancer = pmc.cancer_id AND sr.result = 'progression')

UNION ALL

SELECT
    'Cohort 4' AS cohort_name,
    CASE
        WHEN NOT EXISTS (SELECT 1 FROM molecular_finding mf WHERE mf.cancer = (SELECT cancer_id FROM PatientMainCancer) AND mf.name = 'PD-L1')
        THEN 'Requires PD-L1 TPS >=1%'
        ELSE NULL
    END AS required_details
FROM PatientMainCancer pmc
WHERE
    -- General NSCLC
    pmc.primary_site = 'lung' AND (pmc.histology IS NOT NULL AND LOWER(pmc.histology) NOT LIKE 'small cell%')
    -- General Driver Mutation Exclusion
    AND NOT EXISTS (
        SELECT 1 FROM molecular_finding mf_drv
        WHERE mf_drv.cancer = pmc.cancer_id
        AND mf_drv.name IN ('EGFR', 'ALK', 'ROS1', 'BRAF', 'KRAS', 'MET', 'RET', 'HER2', 'NTRK1', 'NTRK2', 'NTRK3')
        AND mf_drv.finding_type IN ('mutation_or_deletion', 'fusion_or_rearrangement')
        AND mf_drv.result = 'positive'
    )
    -- General Brain Mets Exclusion
    AND (pmc.has_active_brain_mets IS FALSE)

    -- General Cohort Criteria: Stage
    AND pmc.current_extent IN ('locoregional', 'metastatic')
    -- Cohort Specific: ECOG
    AND pmc.ecog_ps <= 2
    -- Cohort Specific: Measurable Disease
    AND pmc.current_extent != 'no_evidence_of_disease'
    -- Cohort Specific: Not candidates for chemotherapy as first-line treatment for advanced/metastatic NSCLC
    AND NOT EXISTS (
        SELECT 1
        FROM systemic_regimen sr
        JOIN systemic_drug sd ON sr.id = sd.regimen
        WHERE sr.cancer = pmc.cancer_id
          AND sr.treatment_setting IN ('advanced', 'metastatic')
          AND sr.sort_order = 1
          AND (sd.atc_third_level_code IN ('L01A', 'L01B', 'L01C', 'L01D') OR sd.atc_fourth_level_code IN ('L01XA', 'L01XB'))
    )
    -- Cohort Specific: PD-L1 TPS >=1%
    AND (
        NOT EXISTS (SELECT 1 FROM molecular_finding mf_pdl1 WHERE mf_pdl1.cancer = pmc.cancer_id AND mf_pdl1.name = 'PD-L1' AND CAST(mf_pdl1.result AS INTEGER) < 1)
    )

UNION ALL

SELECT
    'Cohort 5' AS cohort_name,
    NULL AS required_details -- No specific molecular markers required by name for Cohort 5 beyond general exclusions
FROM PatientMainCancer pmc
WHERE
    -- General NSCLC
    pmc.primary_site = 'lung' AND (pmc.histology IS NOT NULL AND LOWER(pmc.histology) NOT LIKE 'small cell%')
    -- General Driver Mutation Exclusion
    AND NOT EXISTS (
        SELECT 1 FROM molecular_finding mf_drv
        WHERE mf_drv.cancer = pmc.cancer_id
        AND mf_drv.name IN ('EGFR', 'ALK', 'ROS1', 'BRAF', 'KRAS', 'MET', 'RET', 'HER2', 'NTRK1', 'NTRK2', 'NTRK3')
        AND mf_drv.finding_type IN ('mutation_or_deletion', 'fusion_or_rearrangement')
        AND mf_drv.result = 'positive'
    )
    -- General Brain Mets Exclusion (stricter for C5, but pmc.has_active_brain_mets IS FALSE is the check based on available data)
    AND (pmc.has_active_brain_mets IS FALSE)

    -- Cohort Specific: Stage (Unresectable Stage III)
    AND pmc.current_extent = 'locoregional'
    -- Cohort Specific: ECOG
    AND pmc.ecog_ps <= 2
    -- Cohort Specific: Measurable disease NOT required.
    -- Cohort Specific: Undergone chemoradiotherapy
    AND EXISTS (
        SELECT 1
        FROM systemic_regimen sr
        JOIN systemic_drug sd ON sr.id = sd.regimen
        WHERE sr.cancer = pmc.cancer_id
          AND sr.treatment_setting = 'concurrent_radiation'
          AND (sd.atc_third_level_code IN ('L01A', 'L01B', 'L01C', 'L01D') OR sd.atc_fourth_level_code IN ('L01XA', 'L01XB'))
    )

UNION ALL

SELECT
    'Cohort 6' AS cohort_name,
    NULL AS required_details
FROM PatientMainCancer pmc
WHERE
    -- General NSCLC
    pmc.primary_site = 'lung' AND (pmc.histology IS NOT NULL AND LOWER(pmc.histology) NOT LIKE 'small cell%')
    -- General Driver Mutation Exclusion
    AND NOT EXISTS (
        SELECT 1 FROM molecular_finding mf_drv
        WHERE mf_drv.cancer = pmc.cancer_id
        AND mf_drv.name IN ('EGFR', 'ALK', 'ROS1', 'BRAF', 'KRAS', 'MET', 'RET', 'HER2', 'NTRK1', 'NTRK2', 'NTRK3')
        AND mf_drv.finding_type IN ('mutation_or_deletion', 'fusion_or_rearrangement')
        AND mf_drv.result = 'positive'
    )
    -- General Brain Mets Exclusion (stricter for C6, but pmc.has_active_brain_mets IS FALSE is the check)
    AND (pmc.has_active_brain_mets IS FALSE)

    -- Cohort Specific: Stage (Resectable Stage II and Stage III NSCLC) - proxied by localized/locoregional
    AND pmc.current_extent IN ('localized', 'locoregional')
    -- General Cohort Criteria: ECOG
    AND pmc.ecog_ps <= 1
    -- Cohort Specific: Measurable Disease
    AND pmc.current_extent != 'no_evidence_of_disease'
    -- Cohort Specific: NSCLC must be considered technically and medically resectable (assumed if stage matches and other criteria met)

UNION ALL

SELECT
    'Cohort 7' AS cohort_name,
    NULL AS required_details
FROM PatientMainCancer pmc
WHERE
    -- General NSCLC
    pmc.primary_site = 'lung' AND (pmc.histology IS NOT NULL AND LOWER(pmc.histology) NOT LIKE 'small cell%')
    -- General Driver Mutation Exclusion
    AND NOT EXISTS (
        SELECT 1 FROM molecular_finding mf_drv
        WHERE mf_drv.cancer = pmc.cancer_id
        AND mf_drv.name IN ('EGFR', 'ALK', 'ROS1', 'BRAF', 'KRAS', 'MET', 'RET', 'HER2', 'NTRK1', 'NTRK2', 'NTRK3')
        AND mf_drv.finding_type IN ('mutation_or_deletion', 'fusion_or_rearrangement')
        AND mf_drv.result = 'positive'
    )
    -- General Brain Mets Exclusion
    AND (pmc.has_active_brain_mets IS FALSE)

    -- General Cohort Criteria: Stage
    AND pmc.current_extent IN ('locoregional', 'metastatic')
    -- General Cohort Criteria: ECOG
    AND pmc.ecog_ps <= 1
    -- Cohort Specific: Measurable Disease
    AND pmc.current_extent != 'no_evidence_of_disease'
    -- Cohort Specific: Prior therapy (PD-1/L1 and Platinum)
    AND EXISTS (SELECT 1 FROM systemic_drug sd JOIN systemic_regimen sr ON sd.regimen = sr.id WHERE sr.cancer = pmc.cancer_id AND sd.atc_fourth_level_code = 'L01FF')
    AND EXISTS (SELECT 1 FROM systemic_drug sd JOIN systemic_regimen sr ON sd.regimen = sr.id WHERE sr.cancer = pmc.cancer_id AND sd.atc_fourth_level_code = 'L01XA')
    -- Cohort Specific: Must present with progressive disease
    AND EXISTS (SELECT 1 FROM systemic_regimen sr WHERE sr.cancer = pmc.cancer_id AND sr.result = 'progression')

UNION ALL

SELECT
    'Cohort 8' AS cohort_name,
    NULL AS required_details
FROM PatientMainCancer pmc
WHERE
    -- General NSCLC
    pmc.primary_site = 'lung' AND (pmc.histology IS NOT NULL AND LOWER(pmc.histology) NOT LIKE 'small cell%')
    -- General Driver Mutation Exclusion
    AND NOT EXISTS (
        SELECT 1 FROM molecular_finding mf_drv
        WHERE mf_drv.cancer = pmc.cancer_id
        AND mf_drv.name IN ('EGFR', 'ALK', 'ROS1', 'BRAF', 'KRAS', 'MET', 'RET', 'HER2', 'NTRK1', 'NTRK2', 'NTRK3')
        AND mf_drv.finding_type IN ('mutation_or_deletion', 'fusion_or_rearrangement')
        AND mf_drv.result = 'positive'
    )
    -- General Brain Mets Exclusion
    AND (pmc.has_active_brain_mets IS FALSE)

    -- General Cohort Criteria: Stage
    AND pmc.current_extent IN ('locoregional', 'metastatic')
    -- General Cohort Criteria: ECOG
    AND pmc.ecog_ps <= 1
    -- Cohort Specific: Measurable Disease
    AND pmc.current_extent != 'no_evidence_of_disease'
    -- Cohort Specific: Prior therapy (PD-1/L1 and Platinum)
    AND EXISTS (SELECT 1 FROM systemic_drug sd JOIN systemic_regimen sr ON sd.regimen = sr.id WHERE sr.cancer = pmc.cancer_id AND sd.atc_fourth_level_code = 'L01FF')
    AND EXISTS (SELECT 1 FROM systemic_drug sd JOIN systemic_regimen sr ON sd.regimen = sr.id WHERE sr.cancer = pmc.cancer_id AND sd.atc_fourth_level_code = 'L01XA')
    -- Cohort Specific: Must present with progressive disease
    AND EXISTS (SELECT 1 FROM systemic_regimen sr WHERE sr.cancer = pmc.cancer_id AND sr.result = 'progression')

UNION ALL

SELECT
    'Cohort 9' AS cohort_name,
    NULL AS required_details
FROM PatientMainCancer pmc
WHERE
    -- General NSCLC
    pmc.primary_site = 'lung' AND (pmc.histology IS NOT NULL AND LOWER(pmc.histology) NOT LIKE 'small cell%')
    -- General Driver Mutation Exclusion
    AND NOT EXISTS (
        SELECT 1 FROM molecular_finding mf_drv
        WHERE mf_drv.cancer = pmc.cancer_id
        AND mf_drv.name IN ('EGFR', 'ALK', 'ROS1', 'BRAF', 'KRAS', 'MET', 'RET', 'HER2', 'NTRK1', 'NTRK2', 'NTRK3')
        AND mf_drv.finding_type IN ('mutation_or_deletion', 'fusion_or_rearrangement')
        AND mf_drv.result = 'positive'
    )
    -- General Brain Mets Exclusion
    AND (pmc.has_active_brain_mets IS FALSE)

    -- General Cohort Criteria: Stage
    AND pmc.current_extent IN ('locoregional', 'metastatic')
    -- General Cohort Criteria: ECOG
    AND pmc.ecog_ps <= 1
    -- Cohort Specific: Measurable Disease
    AND pmc.current_extent != 'no_evidence_of_disease'
    -- Cohort Specific: Prior therapy (PD-1/L1 and Platinum)
    AND EXISTS (SELECT 1 FROM systemic_drug sd JOIN systemic_regimen sr ON sd.regimen = sr.id WHERE sr.cancer = pmc.cancer_id AND sd.atc_fourth_level_code = 'L01FF')
    AND EXISTS (SELECT 1 FROM systemic_drug sd JOIN systemic_regimen sr ON sd.regimen = sr.id WHERE sr.cancer = pmc.cancer_id AND sd.atc_fourth_level_code = 'L01XA')
    -- Cohort Specific: Must present with progressive disease
    AND EXISTS (SELECT 1 FROM systemic_regimen sr WHERE sr.cancer = pmc.cancer_id AND sr.result = 'progression');
```
